# Singing for lung health following completion of pulmonary rehabilitation - feasibility of a randomised controlled trial

**DOI:** 10.1101/2025.02.10.25321748

**Authors:** Adam Lewis, Peter Jung, Parris J. Williams, Jennifer Steinmann, Karen Ingram, Noah Longley, Puja Trivedi, Stuart Clarke, Helen Lammin, George Edwards, Maria Koulopolou, Arun Sureshkumar, Anna Moore, Paul E. Pfeffer, Leanne Reardon, Kim Sorley, Jarvis Kenman, Brendan DeLuca, Michelle Maguire, Laura-Jane E. Smith, Sarah Elkin, Adam Lound, Laura Moth, Penelope Rickman, Sharon Alexander, Natasha Lohan, Emily Garsin, Susan Young, Amanda Harris, Rosie Watters, Cleo Lane, Claire M Nolan, Joy Conway, William D-C Man, Winston Banya, Nana Anokye, Keir E.J. Philip, Phoene Cave, Nicholas S. Hopkinson

**Affiliations:** School of Health Sciences, University of Southampton, UK; College of Health Medicine and Life Sciences, Department of Health Sciences, Brunel University London, UK; Royal Brompton and Harefield Hospitals, Guy’s and St Thomas’ NHS Foundation Trust, UK; National Heart and Lung Institute, Imperial College London, UK; King’s Centre for Lung Health, Faculty of Life Sciences & Medicine, King’s College London; Kings College Hospital NHS Foundation Trust, UK; Barts Health NHS Trust, UK; South London Clinical Research Network, UK; Imperial College Healthcare NHS Trust, UK; Belfast Health and Social Care Trust, UK; The Musical Breath Ltd, UK

## Abstract

**Background:** Pulmonary rehabilitation(PR) is a highly effective intervention for people with chronic respiratory disease, however it is not known how best to sustain its benefits. Clinical trials are needed to establish if participation in Singing for Lung Health(SLH) groups following PR will improve health-related quality-of-life, healthcare utilisation and exercise capacity compared to usual care. A feasibility study would help to guide development of these.

**Methods:** In a multi-centre, mixed-methods randomised controlled feasibility trial, PR participants at 4 sites, were pre-screened at baseline assessment. An SLH taster session was included routinely as part of the PR programmes. Eligible PR completers were invited to take part in the trial and randomised to usual care or a 12-week SLH course. Feasibility outcomes included recruitment rate, intervention compliance (at least 8/12 sessions) and completeness of data collection including symptom questionnaires, walk tests and physical activity monitoring. Interviews with participants and study personnel about their experience and views of the study were subjected to thematic analysis.

**Results:** Between October 2022 and November 2023, 1311 patients were assessed to start PR, 838 completed. Of those completing, 243 were ineligible to take part, (predominantly due to vaccination status and other primary PR diagnoses), and 531 declined. 64 people (33 female, mean(SD) age 69(12), 41 ethnically white, COPD/asthma/interstitial lung disease/bronchiectasis n=33/16/9/6) were recruited with 30(93.8%) SLH and 29(90.6%) controls completing the study. 20(62.5%) of the SLH group completed at least 8/12 SLH sessions. There was enthusiasm for a definitive trial from participants, clinicians and singing group leaders perspectives based on positive experiences of trial involvement. Improvements to recruitment strategy, intervention structure, outcome measures and staffing were suggested.

**Conclusion:** A definitive RCT of SLH post-PR appears feasible, with acceptable uptake and completion rates.

**Trial Registration:** ISRCTN11056049

**What is already known on this topic?:** Singing for Lung Health(SLH) has previously been shown to improve health-related quality-of-life for people with Chronic Obstructive Pulmonary Disease. Pulmonary Rehabilitation is a gold standard intervention, but it is not known whether SLH groups can be delivered as a maintenance programme after PR completion, or whether an RCT comparing this approach to usual care is feasible.

**What this study adds:** It is feasible to conduct an RCT investigating the clinical and cost effectiveness of a 12-week SLH post PR maintenance programme compared with usual care.

**How this might affect research, practice or policy:** This study will inform the design and delivery of a definitive RCT. The feasibility methodology used in this study can be applied to other creative health interventions which may be considered as maintenance options post-PR.

## INTRODUCTION

Respiratory diseases are among the leading causes of morbidity and mortality globally and affect 1 in 5 people in the UK(2) costing approximately £11 billion per year(3). Despite optimal treatment, many individuals remain disabled by physical impacts to health-related quality of life (4), and social isolation(5). Pulmonary Rehabilitation is a complex intervention consisting of group-based education and exercise, primarily aimed at people living with activity limiting breathlessness. 6 to 8 week programmes of Pulmonary Rehabilitation (PR) are well established with a Grade A evidence base, improving breathlessness, quality of life and exercise tolerance(6) and individualised exercise programme plans should be provided to patients after PR completion (7)(8). The benefits of PR decline over time. Evidence suggests that PR maintenance programmes can reduce the risk of respiratory-related hospital admission, exacerbations and mortality, but further high-quality RCTs are needed(9). Additional approaches are needed to extend the benefits achieved in PR, especially ones which can build communities of social support for patients with chronic respiratory disease (CRD) who experience loneliness, social disengagement and isolation with negative consequences to their health(10).

Singing for Lung Health (SLH) has potential to be offered as a choice of maintenance activity. It is a creative health activity which addresses physical, mental and social needs for patients with respiratory disease(11–15). SLH groups are led by individuals who have received specific training to deliver group singing sessions for people with respiratory disease. If delivered after PR completion, SLH participation could address the need for a maintenance strategy at a critical point where patients have gained significant health benefit and a sense of belonging in a group environment(16). A recent study reported that PR completers who participated in singing as exercise during PR were more likely to report improved breathing control 4.7 years later, compared to those who performed resistance and endurance-based exercises within PR(17).

The aim of this study was to investigate the feasibility of conducting an RCT evaluating the impact of 12 weeks of SLH exercises compared to usual care for people with chronic respiratory diseases following completion of PR. We wanted to test components of a randomised control design to optimise a definitive clinical and cost effectiveness study design.

## METHODS

This study was prospectively registered (ISRCTN11056049) 17/09/21 and approved by Hampshire Research Ethics Committee B reference 21/SC/0240 IRAS number: 293580. 01/10/2021.

We conducted a parallel group, assessor-blind, randomised controlled feasibility study with a nested qualitative study, to address the question; In people with Chronic Obstructive Pulmonary Disease (COPD), asthma, interstitial lung disease (ILD) or bronchiectasis who have completed PR, is an RCT comparing the effect of once weekly SLH to usual care alone feasible for participants to complete Singing for Lung Health Groups?

### Participant enrolment

Four PR centres following British Thoracic Society PR Quality Standards, who had SLH groups in their locality took part in the study, with patients enrolled between October 2022 and November 2023. Following their baseline PR assessment, patients received a brief information leaflet describing the study. The PR programmes were also requested to include a SLH ‘taster’ session as part of the education programme. This included a brief PowerPoint introducing the trial and a demonstration of SLH exercises facilitated by the singing leader, which PR participants and clinicians were encouraged to join in with. Participants were provided participant information sheets following this. Potential participants were invited to attend screening and assessment to take part in the clinical trial no later than a month after their discharge appointment from PR.

### Eligibility Criteria

Participants were eligible for the study if they were at least 18 years old, had received three SARS-CoV-2 vaccines, an influenza and pneumococcus vaccine, clinically diagnosed with COPD, asthma, bronchiectasis or ILD, had stable respiratory health within last 4 weeks (no exacerbations), completed at least eight sessions of PR and were able to provide informed consent. Participants were excluded if they had previously attended SLH group sessions, regularly participated in any other singing group activity, or if they had a life-limiting co-morbidity such as a terminal cancer diagnosis.

### Outcomes

The study investigated a range of measures around recruitment and retention that might determine the feasibility of a definitive clinical trial. The primary feasibility outcome was completion of 12 weeks of SLH group classes. We hypothesised that 60% of individuals who were enrolled onto the study after completing PR, and randomised to the SLH arm, would complete SLH (at least 8 out of 12 sessions). Other secondary feasibility, mechanistic and clinical outcomes were assessed by a face to face, blinded researcher assessment at 12 weeks and questionnaire survey at 24 weeks, including home exercise diary collection (feasibility), physical activity monitoring using McRoberts Dynaport movemonitors (feasibility and mechanistic), adverse events (collected by participant self-report, at a 6 week follow up telephone call, 12 week follow-up and independent telephone contact made by participants) (clinical), and qualitative interview data (feasibility and mechanistic). A list of secondary outcomes is below in Table 1 which are written in chronological order, at which point during the study they were collected and categorised into type of outcome:

**Table 1:**
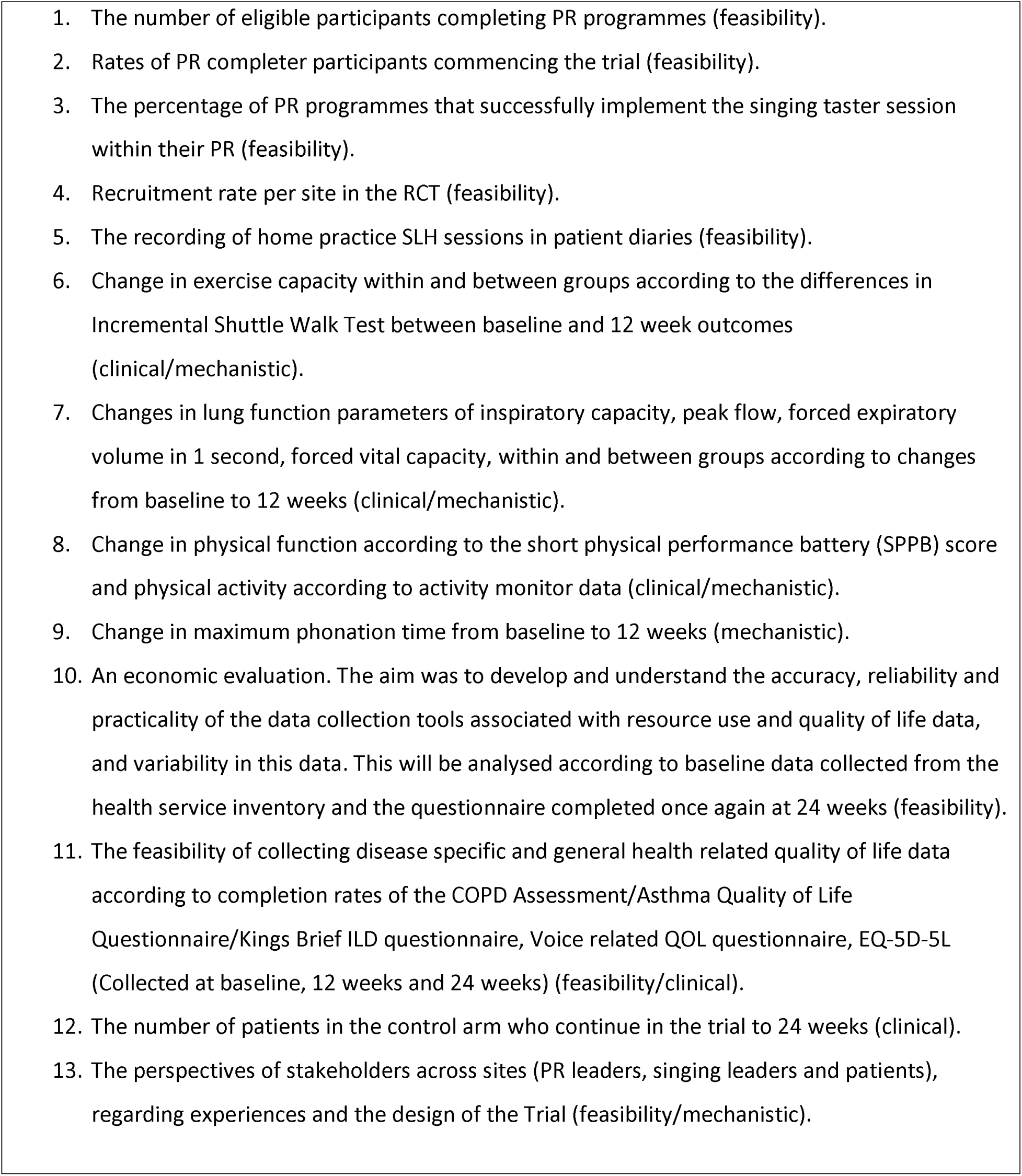
Study outcomes.

### Randomisation

Assessor blind randomisation was on a 1:1 basis based on a REDCap software random sequence generated by the Trial statistician with random block sizes.

### Study arms

#### Usual Care

All study participants were prescribed home exercise advice as a continuation of the clinical PR programme. This included resistance and endurance training including exercises such as bicep curls, lateral shoulder raises, push ups against the wall, squats, sit-to-stands and step-ups using the British Lung Foundation home exercise diary. Individuals were further advised to record their total minutes of outdoor walking each day.

#### Intervention

Additionally, participants randomised to the intervention arm took part in once weekly SLH group sessions for 12 weeks. Singing leaders were all trained by PC who is a world leading expert in SLH delivery. PC coordinated and supported singing leaders throughout the trial. Singing is a complex intervention, involving postural and breathing support, and vocal technique. SLH differs from participation in more generic singing activities by its focus on improving breath control and posture in relation to respiratory disease. A typical 60-minute class includes physical warm-ups, breathing exercises, vocal warm-ups, songs and a cool down/relaxation. Physical warm-ups use body mobilisation and simple exercises as well as using imaginative play, action songs and body percussion. Breathing exercises focus on optimal use of supporting musculature during inhalation and exhalation as well as systematically extending the outbreath. Vocal exercises include unvoiced and voiced fricatives introducing the additional resistance of both a semi occluded vocal tract and vocal fold closure moving from unconscious tidal breathing to a consciously voiced exhale. Finally, ‘Primal sounds’ such as Hey, Ho, Ha, etc., were introduced to engage vocal mechanism and support(18). Such SLH have been used successfully in previous studies (19–21). Daily 15-20 minute practice exercise sessions at home were performed using the ‘Singing for Breathing’ CD given to the participants at the start of their participation in the singing group. The CDs were also available in digital format https://www.themusicalbreath.com/2021/05/13/singing-for-breathing-cd-downloads/ depending on participant preference. The frequency of practice sessions was recorded using home exercise diaries.

This paper has been written in accordance with the Consort 2010 statement extension to pilot and feasibility studies (22).

Both study arms received a phone call follow up at 6 weeks to ask about adverse events and home exercise activity.

### Patient and Public Involvement

PRi and SA were PPI leads in the grant development process and continued to be integral to the study, participating in steering committee meetings throughout the trial, reviewing documents, and supporting dissemination. They both live with chronic respiratory disease. Pri has completed PR and SA has significant experience of SLH.

### Statistical Analysis

Sixty-four participants is a suitable sample size for intervention group feasibility studies(23). If we identified 64 eligible subjects, we estimated a participation rate of 80% to within a 95% confidence interval of +/-10%. We therefore aimed to reach 50 completers of the trial. We were guided in the choice of the sample based on the recommendation of Richard Hooper’s, “Justifying sample size for a feasibility” guidance provided by the Research Design Service London and the audit undertaken by Sophie Bellingham et al (24) who concluded that across feasibility and pilot studies, the median sample size per arm was 36 (range 10 to 300) for trials with a dichotomous endpoint and 30 (range 8 to 114) for trials with a continuous endpoint.

Exploratory analysis of change in outcomes between baseline and the 12-week endpoint was calculated and compared between intervention and study arms using independent two-tailed t-test using intention to treat analysis. Missing change was imputed at zero change. Per protocol sensitivity analysis was performed including those in the intervention arm that adhered to the design. Further sensitivity analysis was performed excluding disease groups with less than 10 participants to assess whether these contributed to greater variability in outcome. Sub analysis was performed in the intervention arm meeting the primary endpoint of adherence to test the change from baseline to 12 weeks using paired two-tailed t-test. The time within metabolic equivalent categories was expressed as a percentage of total time in sedentary, moderate and vigorously active categories.

Standardised effect sizes (d) for change in outcomes at 12-week endpoint between arms were calculated using G*Power. Sample size estimates were calculated to power a prospective trial design at 90% with a 1:1 ratio (alpha 0.05, 1-beta 0.9, two-tailed). Specific sample sizes for d>0.4 were calculated.

### Qualitative assessment

Semi-structured interviews with 10 SLH group participants (completer and non-completers), 5 PR leads and 5 singing leaders was performed to explore their experiences of the trial. Interviews were performed via MS Team/Zoom or by telephone dependant on participant choice. ALe performed the interviews and analysis. He is experienced in both qualitative research and pulmonary rehabilitation. Interviews were transcribed verbatim. Considering the interviewer’s experience being viewed as valuable in the interpretation of findings reflexive thematic analysis(25) was performed. Qualitative methodological considerations were also considered in the analysis considering findings will be acted upon in the design of a future definitive trial(26). The topic guide is provided in the supplementary appendices. The qualitative data aimed to provide a greater depth of understanding and context to quantitative outcomes collected in addition to novel insights into experiences of trial participation and perceptions on the study design.

## RESULTS

1311 patients were screened for eligibility, of whom 595 were eligible. The main reason of ineligibility was non-completion of PR (n=473). 64 patients entered the trial, of whom 59 completed their follow-up 12 week re-assessment. Further details on recruitment and study flow are in figure 1. Participants in the control and intervention groups were well matched at baseline as seen in table 1.

**Figure 1:**
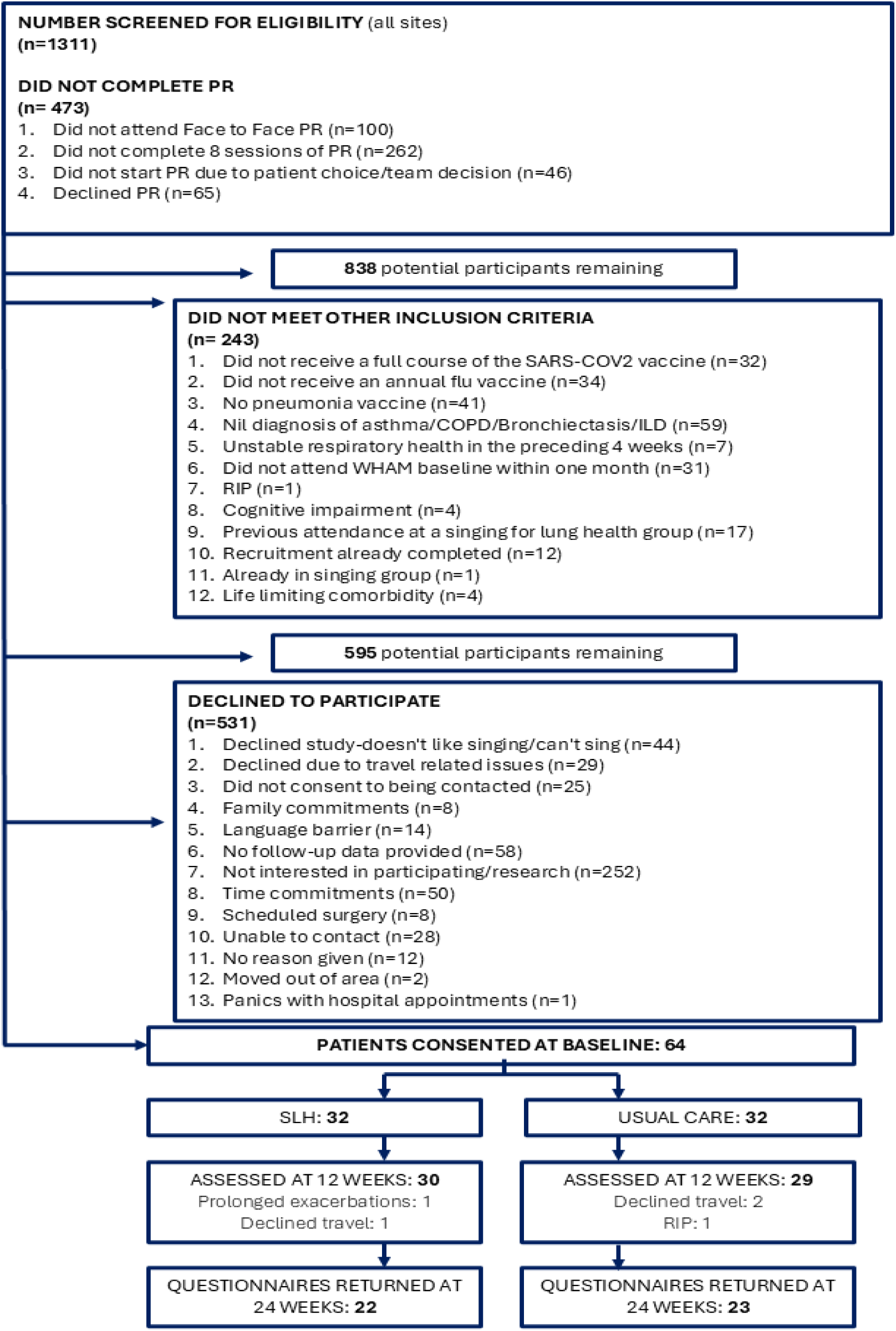
Participant recruitment screening and study completion flow chart.

**Table 1.**
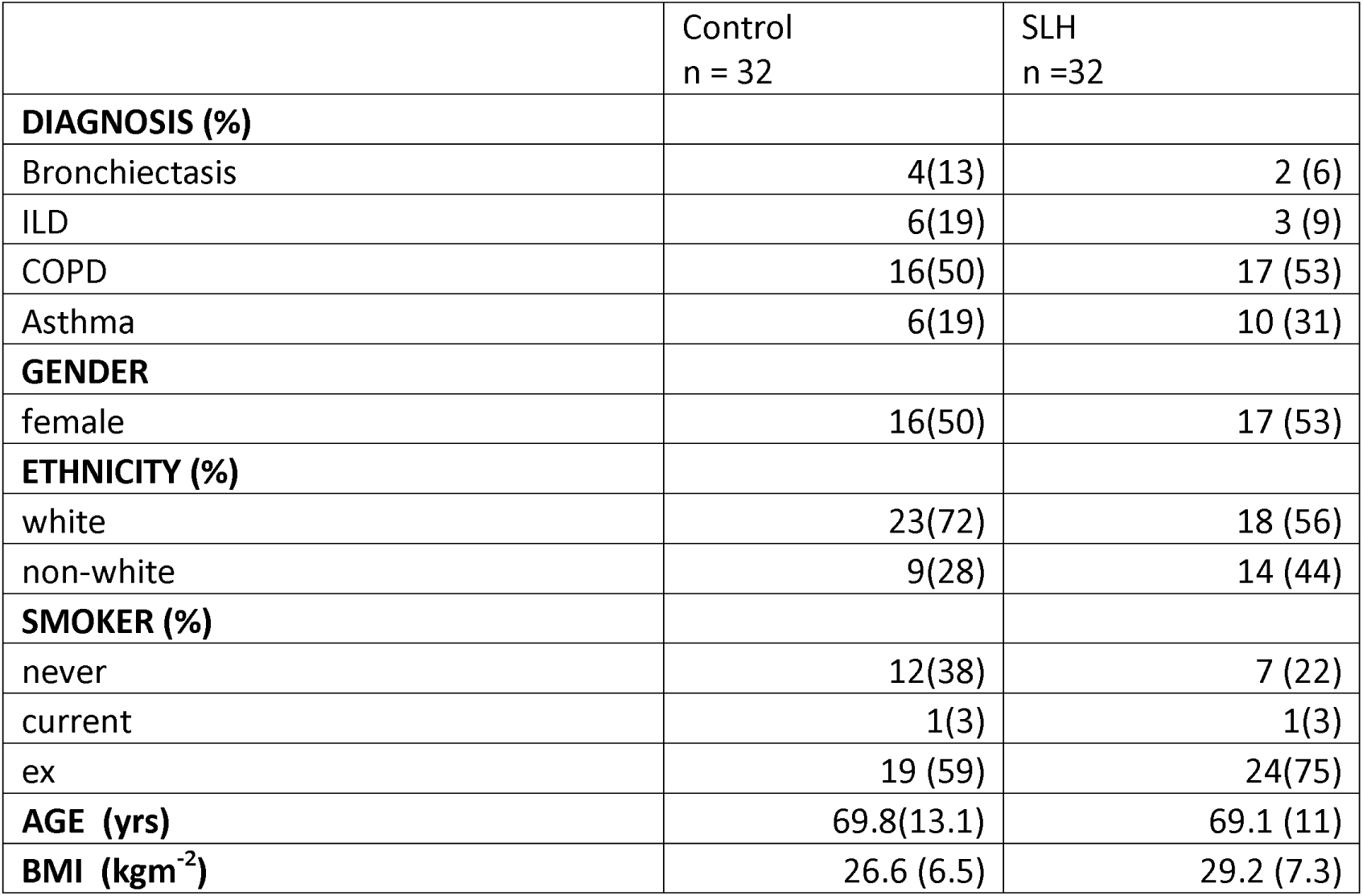
Baseline Demographics. ILD= interstitial lung disease, COPD = chronic obstructive pulmonary disease, BMI = Body Mass Index, values arem ean(SD) or n(%) SD = Standard Deviation

|  | Control<br>n = 32 | SLH<br>n =32 |
| --- | --- | --- |
| **DIAGNOSIS (%)** |  |  |
| Bronchiectasis | 4(13) | 2 (6) |
| ILD | 6(19) | 3 (9) |
| COPD | 16(50) | 17 (53) |
| Asthma | 6(19) | 10 (31) |
| **GENDER** |  |  |
| female | 16(50) | 17 (53) |
| **ETHNICITY (%)** |  |  |
| white | 23(72) | 18 (56) |
| non-white | 9(28) | 14 (44) |
| **SMOKER (%)** |  |  |
| never | 12(38) | 7 (22) |
| current | 1(3) | 1(3) |
| ex | 19 (59) | 24(75) |
| **AGE (yrs)** | 69.8(13.1) | 69.1 (11) |
| **BMI (kgm<sup>-2</sup>)** | 26.6 (6.5) | 29.2 (7.3) |

**Table 2.** Patient outcome measures at Baseline. Unless stated otherwise numbers are represented as mean (standard deviation). FEV1/FVC = Forced expiratory volume in 1^st^ second/Forced Vital Capacity, pFEV1 = percentage of Forced Expiratory Volume in 1^st^ second, pFVC = percentage of predicted forced vital capacity, IC = Inspiratory Capacity, HRQOL = health related quality of life, EQ_VAS = Euroqol visual analogue scale, D12 score = Dyspnoea 12 questionnaire score, VRQOL = voice related quality of life score, MDP_A1 = Multidimensional Dyspnoea Profile unpleasantness, MDP_A2 = Multidimensional Dyspnoea Profile emotional response, MDP_SQ = Multidimensional dyspnoea profile sensory dimension, med = median, ISWT = Incremental Shuttle Walk Test, m = metres, SPPB = Short Physical Performance Battery, EQ_desc = Euroqol descriptive score, EQ_index = Euroqol Index score, MPTS = maximum phonation time in seconds, MET sed% = percentage of time spent in sedentary activity (less than 3 METS), MET mod% = percentage of time spent in moderate activity (3-6 METS), MET act% = percentage of time spent in METS ⇒ 3METS with bout duration 10 minutes allowing interruption of 1 minute. cPPAC = Clinical visit-PROactive Physical Activity in COPD

|  | Control<br>n = 32 | Singing for Lung Health<br>n = 32 |
| --- | --- | --- |
| **Lung Function** |  |  |
| FEV1/FVC | 0.62 (0.19) | 0.64 (0.18) |
| pFVC | 79.69 (22.72) | 82.63 (22.83) |
| pFEV1 | 62.99 (27.22) | 67.27 (26.60) |
| IC | 2.02 (0.64) | 2.03 (0.57) |
| Respiratory rate | 16.8 (4.8) | 16.6 (3.8) |
| **HRQOL** |  |  |
| EQ_vas | 68(18.2) | 65.8 (18.6) |
| EQ_desc (IQR) | 9.5(7-13) | 11 (9-15) |
| EQ_index | 0.673 (0.208) | 0.581 (0.255) |
| D12_score | 10.2 (7.3) | 12.5 (9.3) |
| VRQOL | 83.7 (17.5) | 84.1 (16.6) |
| MDP_A1 | 2.9 (2.2) | 3.4 (2.7) |
| MDP_A2 | 2.5 (2.5) | 2.7 (2.7) |
| MDP_SQ | 2.4 (2.1) | 2.8 (2.8) |
| **OTHER** |  |  |
| Step count (IQR) | 4557 (2586-6453) | 2844 (1851-4802) |
| ISWT (m) (IQR) | 310 (165-455) | 310 (260) |
| SPPB (IQR) | 12 (10-12) | 12 (11-12) |
| EQ_desc (IQR) | 9.5 (7-13) | 11 (8-15) |
| MPTS | 13.2 (6.1) | 15.8 (6.9) |
| MET sed% (med) | 90.40 (87.41-95.31) | 92.73 (88.31) |
| MET mod% (med) | 8.13 (3.47-11.05) | 6.55 (3.66-9.74) |
| MET act% (med) | 1.23 (0.22-2.21) | 0.60 (0 - 1.86) |
| cPPAC total | 65.0 (13.7) | 63.8 (12.6) |
| cPPAC amount | 62.9 (16.1) | 61.5 (12.1) |
| cPPAC difficulty | 67.2 (15.9) | 66.2 (17.3) |

### Recruitment

The target of 64 participants were recruited to the study. 59 participants (92.2%) completed the study at the primary outcome timepoint of 12 weeks and 45 participants (70%) returned data at the 24 week follow up. Five participants were recruited per month on average. Participants were assessed at their baseline WHAM study assessment on average 13.70 days (10.38 SD) after completing PR.

Participant consent per trial site ranged from 3 (2.6% eligible at site) to 43 (19% eligible at site).

### Primary outcome

Twenty out of 32 participants (62.5%) randomised to the SLH + home exercise arm completed eight out of 12 sessions. Of those randomized, there were 2 non-starters of the singing groups; 1 participant was physically assaulted, injured and then unable to attend. The other participant reported it would be too much of a commitment following their initial assessment. There were 10 non-completers with reasons for drop out including: Prolonged chest infection (4), Pneumothorax (not related to intervention) (1), Inappropriate location (2), Travel disruption (storms) (1), caring responsibility (1), travelled abroad for bereavement (1).

23 (72%) of the control group returned postal questionnaire data at 24 weeks and 22 (69%) in the SLH group returned postal questionnaire data at 24 weeks. Therefore, there was not a greater drop out in the control group in this feasibility study.

Exploratory comparative analysis of other clinical outcomes can be found in the online appendices.

Power calculations for a definitive trial can be found in supplementary table S4.

### Singing for Lung Health Taster session integration into PR education programmes

All PR centres integrated SLH taster sessions within their PR programmes.

### Physical Activity Monitoring data

21 participants in both groups (66% of 64) returned paired measurements of the Dynaport Movemonitor and 20 (62.5% of 32) in SLH group and 19 (59.4% of 32) in control group completed paired data for combined PROactive questionnaire and Dynaport MoveMonitor. Table S5 shows pre to post 12-week changes in physical activity data.

### Home exercise diary completion

36/64 participants returned their home exercise diaries. However, of those returned there was little consistency in the way these were completed. Many participants opted to record step counts even though they were advised to record total minutes walked daily.

### Health economic analysis

It was feasible to collect health economic data both by the collection of cost questionnaires at baseline (n = 64/64), 12 weeks (n =59/64) and 24 weeks (45/64), and EQ5D5L data at each time point, although some participants found the cost questionnaire burdensome, particularly recording the number of visits to different healthcare professionals. The number of returns for the economic variables and descriptive statistics can be found in tables S7-10. Data were included of questions that were answered by at least 20 participants. The most popular health economic variable completed was EQ5D5L. 64, 59 and 44 returned completed EQ5D5L questionnaires at baseline, 12 weeks and 6 months respectively.

### Adverse Events

There were 53 adverse events (AEs) and 18 Serious Adverse Events (SAEs) (1 death) in the control group, and 50 AEs and 8 SAEs (0 deaths) in the SLH group. All SAEs were unrelated to the intervention. Total number of adverse events per participant can be found in supplementary table S12.

### Stakeholder perspectives

20 semi-structured interviews were performed. 13 individuals with respiratory conditions were invited to participate in an interview. 1 participant had technical issues and later declined, 1 participant had worsening health and declined, and another participant was leaving the country and declined. 10 participants with respiratory conditions participated in interviews consisting of individuals living with different respiratory conditions, recruited from different PR centres, whether they were randomized to SLH or not and whether they completed a programme or not. Their demographics are in table S11 in the appendices.

5 PR group leaders were interviewed, incorporating 3 out of 4 of the PR centres. Other clinicians were invited from the 4^th^ centre but no response was received following an initial email and reminder. 5 SLH group leaders were interviewed representing all 4 singing group venues.

### Thematic Analysis

Thematic analysis was focused on both understanding the feasibility of the trial design to make suitable adaptions for a future definitive study, whist also understanding the meaning of SLH for participants in the context of pulmonary rehabilitation.

Themes included: Clinical teams were close to capacity; A valuable trial to be involved with; The research process works; Small singing groups; Home exercises not well adhered to; The experience of singing; Social wellbeing an important but missed outcome; Suggested study improvements. Figure 2 shows the thematic map.

**Figure 2:**
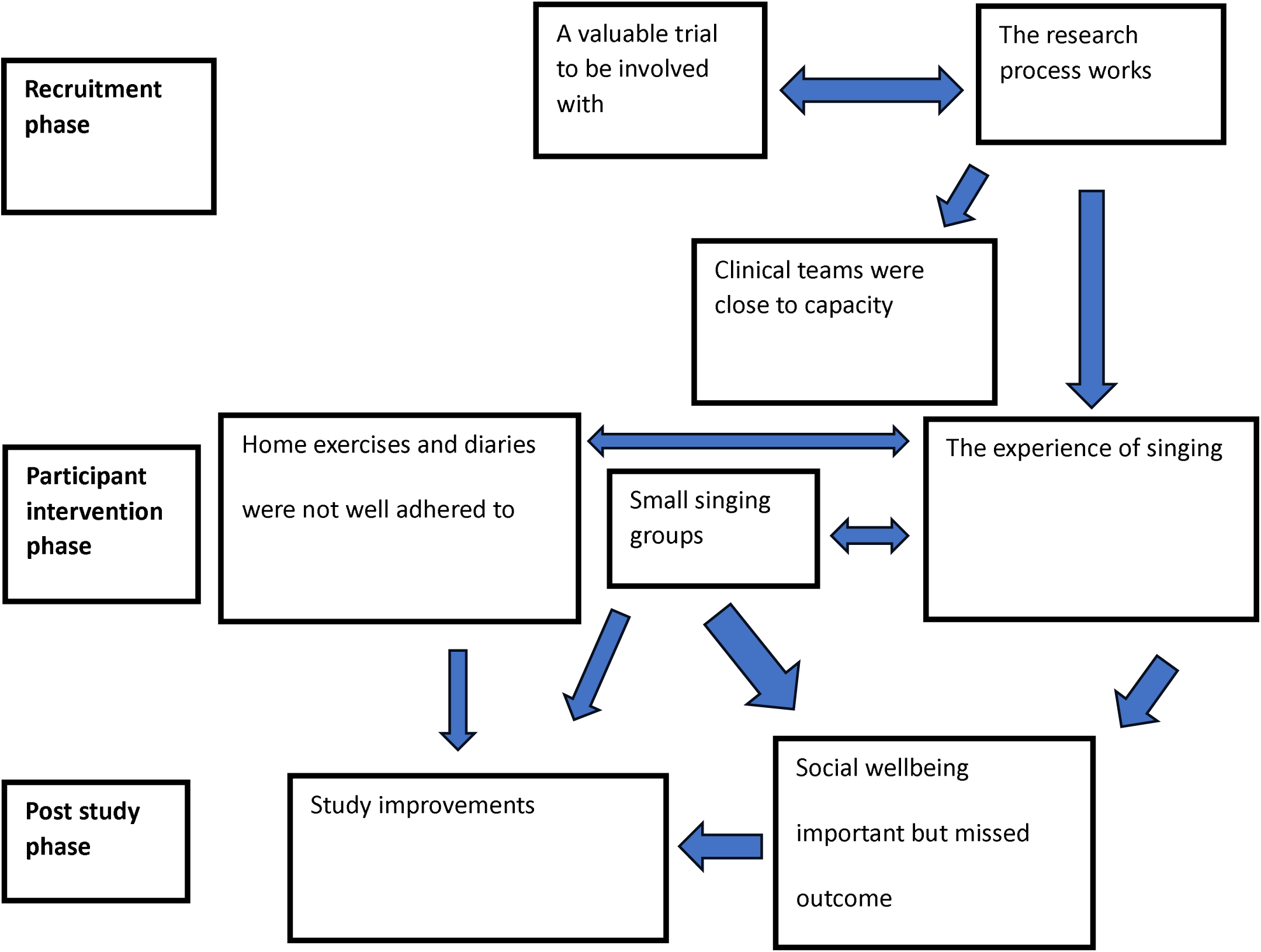
Thematic Map.

### Clinical teams were close to capacity

Multiple teams had staffing difficulties and high turnover during the trial period and were also focused on PR accreditation and other research studies. There was questionable “buy-in” to the trial from some of the clinical teams with a perception from some singing leaders that there was some resistance to an alternative way of working. The WHAM study screening log was not always kept up to date and in some sites communication between the clinical team and singing leaders was sub-optimal. On the other hand, other sites reported being able to integrate the research process into their practice well. It is important to note that clinicians were not funded for their time working on The WHAM Trial.

### A valuable trial to be involved with

Clinicians, patients and singing leaders all felt the trial was valuable. Being involved in research was a good learning opportunity for some clinical staff, and participants felt it was a privilege to be given the opportunity. It was an enjoyable study to be involved with and easy to understand from participant and clinician perspectives. Staff reported they would be keen to be involved in related future studies.

### The research process works

Clinicians were able to discuss the study appropriately with potential participants during PR in a timely manner that did not over burden the PR assessment process. Overall, the WHAM assessment made sense to participants, partly due to the overlap with known PR assessments. Participants reported they understood the importance of randomisation, and the timing of the study after PR was ideal with the singing dose being appropriate. The singing taster sessions were implemented, and they acted as a trigger to discuss the trial and an opportunity to challenge perceptions regarding the purpose of singing within the trial.

### Small singing groups

Singing groups were small across sites during the trial, due to the rolling nature of recruitment, the process of randomization, recruitment rate, and setting up four groups for the study. Participants were expecting to be in larger groups, which reflects those available outside of the trial. Participants and singing leaders felt that although there was value in treating individuals in smaller groups or even on a one-to-one basis, there was huge benefit that came from the group support which was specific and different to that provided in a PR environment.

### Home exercises and diaries were not well adhered to

Prolonged physical activity maintenance post-PR was an acknowledged challenge from both participant and physiotherapist perspectives. Participants could not keep up with the volume of written tasks to do with home exercise diary completion, and the completion of diaries themselves was deliberately not enforced within the SLH sessions. Some felt they needed further instruction on the home exercises at assessment but, in contrast other participants either chose different exercises that fit in with an existing app, or had done similar exercises in the past so knew what to do without the need for a home PR diary.

### The experience of singing

The experience of singing was discussed in terms of how it works, comparisons with PR, focus on breathing control often via the acronym SPLAT (Singers Please Lose Abdominal Tension), and the common belief of “I can’t sing”. Most participants who were interviewed who participated in sessions found benefits in their day to day lives, from enjoyment, feeling they were more motivated to do things, being in control of their breathing, or more comfortable with their condition. However, a participant also noted having to leave the class before the end of a session because she found it difficult, embarrassing and upsetting.

### Social wellbeing important but missed outcome

Participants thought the experience of SLH was inherently social and the environment set up to be pro-social, regarding leaving the house, socializing with others, gaining confidence and self-esteem. The benefits however were also described as intangible and not being able to determine a measurable difference from before the trial.

### Study improvements

Participants, singing leaders and clinicians all suggested study improvements. Participants thought the case report form was too complicated and not all questions were necessary or related to them. It was reported that participants in PR patient identification centre locations are reluctant to leave the local area, and not keen to travel across London for assessments, and so local assessments with clinician research split roles would be advantageous. There were also suggestions made regarding home exercise diary and progression adaptions, with the use of websites, videos, smartwatches and apps needing to be considered.

Supporting quotes aligned to the themes above can be found in Table S6.

Based on the analysis of both qualitative and quantitative data, and further discussions with the study steering group a set of recommendations have been created when considering the design of a future definitive study, in Table 3.

**Table 3.** Recommendations for future study.

|  |
| --- |
| **Recruitment** |
| Trial set up meeting to be included with a group working workshop for PR staff and Singing leaders. Ex PR participants invited to attend. |
| Stratified randomisation according to site |
| Drop stringent exclusion criteria such as the combination of vaccines |
| Use National Respiratory Audit Programme to include sites with higher completion rates of patients |
| Review participant facing research material to focus on breathlessness management more. |
| Expert patient to participate in SLH taster session |
| **Intervention** |
| Start first group once 4 people have been randomised within 3 months of PR completion |
| Digital/app/video options for home exercises would be beneficial |
| Singing leader group practice meetings and reflections embedded within trial supported by the singing coordinator |
| Separate appointment for one-to-one intervention description/demonstration and delivery of home exercises with singing leader before group start. |
| Unblinded allied health professional to deliver home PR intervention with training and demonstration of each PR exercise. |
| Electronic/website diary entry could trigger additional telephone calls from the research team |
| Phased approach to home exercise provision for singing exercises |
| Skeleton referral beneficial for singing leader with information sharing permissions |
| **Patient Outcomes** |
| Social wellbeing and loneliness questionnaire to be considered in assessment battery |
| Singing leaders to have some ownership of SLH home exercise diaries. |
| Video calls (30 mins) for 6 week follow up. |
| Give participants smartwatches that measure sedentary behaviour to vigorous physical activity |
| Change and reduce assessment battery. Simplify the health economic questionnaire. |
| Telephone follow-up of questionnaire data at 24 weeks with face-to-face review as option |
| **Staffing** |
| Clinician researcher Principal Investigator per site needed with local site assessments |
| Repeated in-service trainings with staff about research and SLH throughout the trial |
| Singing for Lung Health education (included as a range of education options) to be included in clinical staff induction. |
| Post doctoral project manager to be funded to coordinate the definitive study. |
| Clinical teams to participate in a local SLH group as part of study set up and knowing local maintenance options (continuing professional development opportunity) |
| Annual budget for each clinical team to be reimbursed for their time working on the study |
| Need main singing practitioners and deputies working in pairs |

## DISCUSSION

This is the first RCT exploring the feasibility of running face-to-face SLH groups after PR completion. This is also the first study to recruit individuals with multiple different chronic respiratory diseases into face-to-face SLH groups. The primary feasibility outcome was achieved in that 20/32 individuals completed at least 8 out of 12 sessions, above the feasibility threshold that we had set. It was possible to recruit participants successfully within a month of completing PR, during which period people are maintaining benefit from PR.

262 out of 1311 (20%) of those who were not eligible for study inclusion was because they did not complete at least 8 sessions of PR. Non-completion of PR is common in clinical practice. Another 100 people did not attend face-to-face PR. For those attending virtual PR, online SLH has been shown to improve quality-of-life for people living with COPD(19). Eligibility criteria were reviewed by a research ethics committee early in COVID-19 in the UK, advising the exclusion of participants without pneumococcal, influenza or covid-19 vaccines which further reduced the numbers of those who could be eligible (107 out of 1311 (8%)). Patients can complete a group session of PR without these vaccines and so it is possible these stringent exclusion criteria could be dropped for a future study.

There was significant heterogeneity in the numbers of eligible participants who were recruited per site. RBHH recruited 19% of eligible patients. This site was where the principal investigator and chief investigator worked, is a flagship national PR centre (re-accredited) and clinicians were used to supporting research. Furthermore, all participants were assessed in this Trust. There were no principal investigators in the other sites, which were patient identification centres. A future study should be funded to have a principal investigator in each site with local participant research assessments.

The participant flow through the study from baseline randomisation to the 12-week primary outcome timepoint, and 6 month follow up indicates feasibility for a future definitive study considering low rates of attrition, and no difference in rates of attrition between groups, indicating willingness of participants to be randomised to the control group. Improvements have been suggested to reduce attrition at the 6 month follow up time point such as offering a face-to-face review or clinician telephone follow-up for questionnaire completion.

The James Lind Alliance breathlessness top 10 research priority setting exercise priority number 2 is focused on how support for breathlessness can be tailored for those from different ethnic and social backgrounds(27). 23 out of 64 (36%) of participants recruited were of non-white ethnicity. We have also collected health economic data suggesting a range of education and occupation backgrounds in this sample, suggesting singing is an intervention of interest for diverse populations with respiratory disease, and one that can certainly be tailored. This is important as demographic variables such as these are not always collected in clinical trials(28).

The completion and return of physical activity monitors at both baseline and paired follow up points in both groups was suboptimal (21/32 in each group) and participants in the qualitative analysis recommended the use of smartwatches and/or apps to record activity in future studies. New monitoring devices would need to be reviewed regarding the accuracy of their recording.

Participants rarely completed the home diaries according to prescription, and interestingly often entered step counts rather than the total minutes of walking activity per day as advised. This may reflect poor patient instruction, design of the diaries, or both. The method of recording home exercises needs optimizing for a future definitive trial.

### Strengths and Limitations

This study indicates multiple components of feasibility regarding the trial design. This is valuable information for a future definitive study. The WHAM study recruited individuals from a range of PR programmes in diverse areas of London, reflecting generalizability of clinical practice and the inclusion of future recruitment sites. Participants, singing leaders, and clinicians all valued the research and recommended a future study following their involvement in The WHAM Study. A range of participant experiences, alongside clinician and singing leaders experiences of the study were obtained in qualitative data, which enabled the generation of themes relevant to multiple stakeholders for a future study. The completion rate is a strength indicating the study was conducted well and participants did not find it too burdensome to return at 12 weeks to a single assessment location.

Many patients needed to be screened to enter the study. Furthermore, during COVID-19 the clinical teams were both focused on working through a waiting list of patients, some focusing on PR accreditation and all services had issues with staffing. The numbers screened needs to be in context with the feasibility outcomes in this specific study. Although the numbers screened were high, future clinical effectiveness studies would commence screening at PR completion. Pulmonary Rehabilitation teams should be provided with funding to support recruitment in a future study, which should also be resourced with the support of a clinical trial network.

## Conclusion

We performed a randomised controlled feasibility study comparing SLH to usual care. The trial showed multiple areas of trial feasibility including recruitment and retention through the trial pre-to-post 12-week outcome completion, and appetite from singing leaders, physiotherapists and patients for a future definitive RCT. Improvements are required regarding the need of a PI in each recruitment site, that has good PR completion rates. The home exercise diaries and health economic questionnaires need to be further adapted, and offering a face-to-face or telephone appointment for 24 week follow-up questionnaire data could be considered. A definitive clinical and cost effectiveness randomised controlled trial of SLH after PR completion is now warranted.

## Data Availability

Data sharing may occur on reasonable request

## Acknowledgements

We would like to thank all the study participants for their study participation. We would also like to thank all other members of clinical teams and supporting admin staff involved in supporting the study. We are grateful for the statistical consultancy services provided by Dr Iain Stewart (www.indigo-sigma.co.uk). Many Thanks to Ed and Elisa Jeffrey for their support during the trial in the production of the singing for lung health taster video.

## Contributors

The study was designed by ALe, NSH, WB, KEJP, ALo, PR, SA, PC, NA, and NSH. Ale and NSH wrote the protocol and obtained ethics approval and authorisations. PJ, PW, JS, KI, NL, PT, SC, HL, GE, MK, AS, LR, KS, JK, BD, MM, contributed to participant recruitment. Ale conducted randomization of patients. PJ, PW, JS, KS completed the quantitative data collection. Ale completed qualitative data collection. The initial statistical analysis plan was devised by WB (medical statistician), with the analysis completed by IDS and Ale. NL, EG, SY, AH, RW, and CL delivered the study intervention. All authors contributed to the study conduct, interpretation, revising the manuscript and agreeing on the final version. Ale and NSH are the guarantors and accept full responsibility for the work and/or the conduct of the study, had access to the data, and controlled the decision to publish.

