## Supplementary Appendices for "Singing for lung health following completion of pulmonary rehabilitation - feasibility of a randomised controlled trial"

University Rd, Highfield

Southampton, UK.

SO17 1BJ

**Funding:**

This project was funded by the National Institute for Health Research (NIHR) under its Research for Patient Benefit (RfPB) Programme (Grant Reference Number NIHR201539). The views expressed are those of the author(s) and not necessarily those of the NIHR or the Department of Health and Social Care.

**COI:** CMN reports grants from the National Institute for Health and Care Research, outside the submitted work. All other authors declare no conflicts of interest.

**Data Sharing Statement:** Data sharing may occur on reasonable request

### Table of Contents

Table S1. Intention to treat analysis	6

Table S2. Per protocol analysis	7

Table S3. Sensitivity Analysis with Asthma and COPD only cohorts	8

Table S4. Power Calculations	9

Table S5: Physical Activity monitoring	10

Table S6. Quotes from participants	11

Table S7: Baseline cohort qualifications	14

Table S8: Baseline Health Economics Data	15

Table S9: 12-week follow-up Health Economics data	17

Table S10: 6-month Health Economics data	19

Table S11. Participant demographics for qualitative interviews	21

Table S12: Adverse events	22

Semi-structured interview guide – Respiratory Participants	24

### Pre-Post 12 week outcomes

Table S1 shows the between group differences at 12 weeks (Intention to treat analysis) in the appendices. Results show there was a significant between group difference at 12 weeks of the CAT score favouring those in the control group (mean diff: 4.16, CI 1.20; 7.12, P value 0.007, effect size 1.) All other outcomes showed no between group difference. Per protocol analyses and disease sensitivity analyses are also provided in the online appendices in tables S2 and S3.

### Table S1. Intention to treat analysis

| **12WEEK BETWEEN GROUP (Intention To Treat) missing imputed at 0** |  |  |  |  |  |  |  |
| --- | --- | --- | --- | --- | --- | --- | --- |
|  | ctrl | n=32 | slh | n=32 | diff | 95%CI | p |
| PRIMARY |  |  |  |  |  |  |  |
| VRQOL_change | -2.3 | (12.1) | -1.6 | (10.7) | 0.7 | -5.0; 6.4 | 0.806 |
| D12_change | 0.16 | (7.08) | -0.56 | (6.52) | -0.72 | -4.12; 2.68 | 0.674 |
| MDPSQ_change | 0.22 | (1.67) | 0.35 | (2.31) | 0.13 | -0.87; 1.14 | 0.792 |
| MDPA1_change | 0.00 | (2.31) | -0.03 | (2.28) | -0.03 | -1.19; 1.11 | 0.957 |
| MDPA2_change | 0.21 | (2.15) | 0.20 | (2.88) | -0.01 | -1.28; 1.25 | 0.982 |
| CAT change | -1.75 | (3.57) | 2.41 | (4.66) | 4.16 | 1.20; 7.12 | 0.007 |
| SECONDARY |  |  |  |  |  |  |  |
| ISWT_change | -18.4 | (72.6) | -4.1 | (67.2) | 14.4 | -20.6; 49.3 | 0.414 |
| SPPB_change | -0.28 | (0.92) | -0.06 | (1.68) | 0.22 | -0.46; 0.90 | 0.522 |
| MPTS_change | -0.10 | (4.66) | -0.37 | (4.75) | -0.28 | -2.63; 2.08 | 0.816 |
| Eqvas_change | -4.4 | (22.1) | -4.4 | (18.6) | -0.06 | -10.3; 10.2 | 0.990 |
| Eqindex_change | 0.022 | (0.149) | 0.023 | (0.204) | 0.001 | -0.089; 0.090 | 0.986 |
| Eqdesc_change | -0.13 | (2.06) | -0.53 | (2.83) | -0.41 | -1.64; 0.83 | 0.514 |
| LFT |  |  |  |  |  |  |  |
| FEV1L_change | -0.08 | (0.23) | -0.05 | (0.19) | 0.03 | -0.07; 0.14 | 0.517 |
| pFEV_change | -2.66 | (9.43) | -5.33 | (19.36) | -2.67 | -10.28; 4.94 | 0.486 |
| FVCL_change | -0.14 | (0.28) | -0.07 | (0.33) | 0.07 | -0.08; 0.23 | 0.332 |
| pFVC_change | -3.78 | (10.20) | -5.65 | (23.09) | -1.87 | -10.79; 7.05 | 0.677 |
| FEV1/FVC_change (%) | 0.05 | (5.67) | 0.57 | (7.55) | 0.52 | -2.82; 3.85 | 0.759 |
| IC_change | -0.03 | (0.28) | -0.03 | (0.21) | 0 | -0.12; 0.13 | 0.948 |
| resprate_change | 1.13 | (15.55) | -0.91 | (3.86) | -2.03 | -7.69; 3.63 | 0.476 |

Table S1 Figure Legend VRQOL = voice related quality of life score, D12 = Dyspnoea 12 questionnaire score, MDP_A1 = Multidimensional Dyspnoea Profile unpleasantness, MDP_A2 = Multidimensional Dyspnoea Profile emotional response, MDP_SQ = Multidimensional dyspnoea profile sensory dimension, CAT = Chronic Obstructive Pulmonary Disease Assessment Test, ISWT = Incremental Shuttle Walk Test, SPPB = Short Physical Performance Battery, MPTS = maximum phonation time in seconds, EQ_vas = Euroqol visual analogue scale, med = median, , m = metres, EQ_desc = Euroqol descriptive score, EQ_index = Euroqol Index score, LFT = Lung Function Tests, FEV1/FVC = Forced expiratory volume in 1^st^ second/Forced Vital Capacity, pFEV1 = percentage of Forced Expiratory Volume in 1^st^ second, pFVC = percentage of forced vital capacity, IC = Inspiratory Capacity, resprate = respiratory rate.

### Table S2. Per protocol analysis

| **12WEEK BETWEEN GROUP (Per Protocol) missing imputed at 0** |  |  |  |  |  |  |  |
| --- | --- | --- | --- | --- | --- | --- | --- |
|  | ctrl | n=32 | slh | n=20 | diff | 95%CI | p |
| PRIMARY |  |  |  |  |  |  |  |
| VRQOL_change | -2.3 | (12.1) | -2.4 | (10.9) | -1.1 | -6.8; 6.5 | 0.973 |
| D12_change | 0.16 | (7.08) | 0.30 | (6.49) | 0.14 | -3.72; 4.00 | 0.941 |
| MDPSQ_change | 0.22 | (1.67) | 0.28 | (2.56) | 0.06 | -1.25; 1.38 | 0.924 |
| MDPA1_change | 0.00 | (2.31) | -0.05 | (2.48) | -0.05 | -1.44; 1.34 | 0.943 |
| MDPA2_change | 0.21 | (2.15) | 0.40 | (2.23) | 0.19 | -1.08; 1.45 | 0.766 |
| SECONDARY |  |  |  |  |  |  |  |
| ISWT_change | -18.4 | (72.6) | 2.0 | (63.0) | 20.4 | -17.9; 58.8 | 0.289 |
| SPPB_change | -0.28 | (0.92) | 0.35 | (1.87) | 0.63 | -0.29; 1.56 | 0.172 |
| MPTS_change | -0.10 | (4.66) | -1.16 | (4.12) | -1.06 | -3.55; 1.42 | 0.394 |
| Eqvas_change | -4.4 | (22.1) | -2.6 | (18.6) | 1.8 | -9.7; 13.3 | 0.757 |
| Eqindex_change | 0.022 | (0.149) | 0.055 | (0.231) | 0.033 | -0.085; 0.152 | 0.571 |
| Eqdesc_change | -0.13 | (2.06) | -1.00 | (3.13) | -0.88 | -2.48; 0.73 | 0.276 |
| LFT |  |  |  |  |  |  |  |
| FEV1L_change | -0.08 | (0.23) | 0.00 | (0.17) | 0.08 | -0.03; 0.19 | 0.140 |
| pFEV_change | -2.66 | (9.43) | -4.41 | (23.47) | -1.74 | 13.12; 9.64 | 0.754 |
| FVCL_change | -0.14 | (0.28) | -0.01 | (0.28) | 0.13 | -0.03; 0.29 | 0.102 |
| pFVC_change | -3.78 | (10.20) | -4.26 | (27.06) | -0.48 | 13.54; 12.59 | 0.941 |
| FEV1/FVC_change | 0.001 | (0.057) | 0.007 | (0.081) | 0.006 | -0.036; 0.048 | 0.770 |
| IC_change | -0.03 | (0.33) | -0.03 | (0.21) | 0.0004 | -0.17; 0.17 | 0.996 |
| resprate_change | 1.24 | (16.35) | -2.29 | (3.42) | -3.08 | -8.84; 2.69 | 0.287 |

Table S2 Figure Legend: VRQOL = voice related quality of life score, D12 = Dyspnoea 12 questionnaire score, MDP_A1 = Multidimensional Dyspnoea Profile unpleasantness, MDP_A2 = Multidimensional Dyspnoea Profile emotional response, MDP_SQ = Multidimensional dyspnoea profile sensory dimension, ISWT = Incremental Shuttle Walk Test, SPPB = Short Physical Performance Battery, MPTS = maximum phonation time in seconds, EQ_vas = Euroqol visual analogue scale, med = median, , m = metres, EQ_desc = Euroqol descriptive score, EQ_index = Euroqol Index score, LFT = Lung Function Tests, FEV1/FVC = Forced expiratory volume in 1^st^ second/Forced Vital Capacity, pFEV1 = percentage of Forced Expiratory Volume in 1^st^ second, pFVC = percentage of forced vital capacity, IC = Inspiratory Capacity, resprate = respiratory rate.

### Table S3. Sensitivity Analysis with Asthma and COPD only cohorts

| **12WEEK BETWEEN GROUP (Per Protocol) missing 0 imputed** |  |  |  |  |  |  |  |
| --- | --- | --- | --- | --- | --- | --- | --- |
|  | ctrl | n=22 | slh | n=17 | diff | 95%CI | p |
| PRIMARY |  |  |  |  |  |  |  |
| VRQOL_change | -1.7 | (9.3) | -2.3 | (11.8) | -0.6 | -7.8; 6.5 | 0.854 |
| D12_change | 0.27 | (7.48) | 0.00 | (6.92) | -0.27 | -4.96; 4.42 | 0.907 |
| MDPSQ_change | 0.11 | (1.89) | -0.17 | (2.41) | -0.28 | -1.73; 1.17 | 0.699 |
| MDPA1_change | -0.45 | (2.20) | -0.12 | (2.69) | 0.34 | -1.30; 1.97 | 0.678 |
| MDPA2_change | 0.15 | (2.56) | 0.21 | (2.29) | 0.07 | -1.51; 1.64 | 0.933 |
| SECONDARY |  |  |  |  |  |  |  |
| ISWT_change | -17.7 | (83.9) | 9.4 | (64.6) | 27.1 | -21.0; 75.2 | 0.289 |
| SPPB_change | -0.23 | (0.87) | 0.65 | (1.80) | 0.87 | -0.11; 1.86 | 0.079 |
| MPTS_change | -0.59 | (4.51) | -1.08 | (4.39) | -0.5 | -3.41; 2.41 | 0.730 |
| Eqvas_change | -3.4 | (24.6) | -1.3 | (20.2) | -2.1 | -16.6; 12.4 | 0.774 |
| Eqindex_change | 0.001 | (0.129) | 0.107 | (0.211) | 0.105 | -0.014; 0.225 | 0.082 |
| Eqdesc_change | 0.18 | (1.84) | -1.47 | (3.16) | -1.65 | -3.43; 0.12 | 0.067 |
| LFT |  |  |  |  |  |  |  |
| FEV1L_change | -0.08 | (0.25) | 0.02 | (0.17) | 0.10 | -0.04; 0.24 | 0.150 |
| pFEV_change | -3.26 | (9.51) | -4.27 | (25.50) | -1.01 | -14.60; 12.57 | 0.878 |
| FVCL_change | -0.18 | (0.29) | 0.06 | (0.22) | 0.24 | 0.07; 0.40 | 0.005 |
| pFVC_change | -5.71 | (8.70) | -2.73 | (28.92) | 2.98 | -12.23; 18.19 | 0.686 |
| FEV1/FVC_change | 0.013 | (0.059) | -0.009 | (0.070) | -0.022 | -0.065; 0.021 | 0.302 |
| IC_change | -0.04 | (0.24) | -0.02 | (0.35) | 0.01 | -0.19; 0.22 | 0.887 |
| resprate_change | 2.59 | (18.32) | -2.00 | (3.50) | -4.59 | -12.86; 3.67 | 0.262 |

Table S3 Figure Legend VRQOL = voice related quality of life score, D12 = Dyspnoea 12 questionnaire score, MDP_A1 = Multidimensional Dyspnoea Profile unpleasantness, MDP_A2 = Multidimensional Dyspnoea Profile emotional response, MDP_SQ = Multidimensional dyspnoea profile sensory dimension, ISWT = Incremental Shuttle Walk Test, SPPB = Short Physical Performance Battery, MPTS = maximum phonation time in seconds, EQ_vas = Euroqol visual analogue scale, med = median, , m = metres, EQ_desc = Euroqol descriptive score, EQ_index = Euroqol Index score, LFT = Lung Function Tests, FEV1/FVC = Forced expiratory volume in 1^st^ second/Forced Vital Capacity, pFEV1 = percentage of Forced Expiratory Volume in 1^st^ second, pFVC = percentage of forced vital capacity, IC = Inspiratory Capacity, resprate = respiratory rate.

### Table S4. Power Calculations

| **Variable** | **ITT (d)** | **ITT (N)** | **PP (d)** | **PP (N)** | **ITT diag sens (d)** | **ITT diag sens (N)** |
| --- | --- | --- | --- | --- | --- | --- |
| VRQOL_change | 0.06 |  | 0.01 |  | 0.06 |  |
| D12_change | 0.11 |  | 0.02 |  | 0.04 |  |
| MDPSQ_change | 0.06 |  | 0.03 |  | 0.13 |  |
| MDPA1_change | 0.01 |  | 0.02 |  | 0.13 |  |
| MDPA2_change | 0.00 |  | 0.09 |  | 0.02 |  |
| ISWT_change | 0.20 |  | 0.30 |  | 0.36 |  |
| SPPB_change | 0.16 |  | 0.43 | 228 | 0.62 | 112 |
| MPTS_change | 0.06 |  | 0.24 |  | 0.11 |  |
| Eqvas_change | 0.00 |  | 0.09 |  | 0.09 |  |
| Eqindex_change | 0.01 |  | 0.17 |  | 0.61 | 116 |
| Eqdesc_change | 0.16 |  | 0.33 |  | 0.64 | 106 |
| FEV1L_change | 0.14 |  | 0.40 | 272 | 0.47 | 194 |
| pFEV_change | 0.18 |  | 0.10 |  | 0.05 |  |
| FVCL_change | 0.23 |  | 0.46 | 198 | 0.91 | 54 |
| pFVC_change | 0.10 |  | 0.02 |  | 0.14 |  |
| FEV1/FVC_change (%) | 0.08 |  | 0.09 |  | 0.34 |  |
| IC_change | 0.00 |  | 0.00 |  | 0.07 |  |
| resprate_change | 0.18 |  | 0.30 |  | 0.35 |  |
| Steps | 0.02 |  | 0.02 |  | 0.05 |  |
| MET sed% change | 0.34 |  | 0.56 | 136 | 0.43 | 228 |
| MET mod% change | 0.33 |  | 0.51 | 166 | 0.36 |  |
| MET act% change | 0.27 |  | 0.80 | 68 | 0.76 | 76 |
| CAT change | 1.00 | 44 | 0.71 | 88 |  |  |

Table S4 Figure Legend: ITT = intention to treat, N = number, d = effect size PP = per protocol, diag sens = including data on those with COPD and Asthma only. Diag VRQOL = voice related quality of life score, D12 = Dyspnoea 12 questionnaire score, MDP_A1 = Multidimensional Dyspnoea Profile unpleasantness, MDP_A2 = Multidimensional Dyspnoea Profile emotional response, MDP_SQ = Multidimensional dyspnoea profile sensory dimension, CAT = Chronic Obstructive Pulmonary Disease Assessment Test, ISWT = Incremental Shuttle Walk Test, SPPB = Short Physical Performance Battery, MPTS = maximum phonation time in seconds, EQ_vas = Euroqol visual analogue scale, med = median, , m = metres, EQ_desc = Euroqol descriptive score, EQ_index = Euroqol Index score, LFT = Lung Function Tests, FEV1/FVC = Forced expiratory volume in 1^st^ second/Forced Vital Capacity, pFEV1 = percentage of Forced Expiratory Volume in 1^st^ second, pFVC = percentage of forced vital capacity, IC = Inspiratory Capacity, resprate = respiratory rate.

### Table S5: Physical Activity monitoring

| **12WEEK BETWEEN GROUP** |  |  |  |  |  |  |  |
| --- | --- | --- | --- | --- | --- | --- | --- |
|  | Ctrl mean | n=21 (SD) | SLH mean | n=21 | Diff (Mean) | 95%CI | p |
| Steps | -618.3 | (2014.3) | -588.6 | (1664.5) | 29.7 | -1122.8; 1182.1 | 0.959 |
| MET sed% change | -0.7 | (15.5) | -7.0 | (21.6) | -6.2 | -18.0; 5.5 | 0.288 |
| MET mod% change | 1.1 | (15.1) | 6.4 | (17.4) | 5.2 | -4.9; 15.4 | 0.304 |
| MET act% change | -0.4 | (1.3) | 0.6 | (5.1) | 1.0 | -1.3; 3.3 | 0.387 |
|  | Ctrl mean | n= 20 (SD) | SLH median | N=19 (IQR) | Diff (Mann-Whitney U) | Z score |  |
| cPPAC Total change | 0.85 | 12.9 | 1 | 10 | 180.5 | -.267 | 0.789 |
| cPPAC Amount change | -3.35 | 14.83 | 0 | 20 | 179 | -.310 | 0.757 |
| cPPAC Difficulty change | 5.05 | 14.84 | 3 | 14 | 158 | -.900 | 0.368 |

Table S5 Figure Legend: MET sed% = percentage of time spent in sedentary activity (less than 3 METS), MET mod% = percentage of time spent in moderate activity (3-6 METS), MET act% = percentage of time spent in METS => 3METS with bout duration 10 minutes allowing interruption of 1 minute. cPPAC = Clinical visit-PROactive Physical Activity in COPD

### Table S6. Quotes from participants

| **Theme** | **Example quotes** |
| --- | --- |
| Clinical teams were close to capacity | We did that explaining more about the study to the patients, and so when it came to the data collection point, like when staffing was a little bit difficult for that time with us, you know during that period, we found it a little bit difficult to collect that data and report the data and send, send the data to the research team.  P11 page P4 line 134 (physiotherapist)  And I just think in that pulmonary rehab space there was, there was a lot happening at that time, which it probably always will be and it's just trying to keep it (WHAM) on top of the agenda for making sure that, you know, you do a little training session, perhaps when people, people are inducted.  P15 page 42 line 1446 (physiotherapist)  That was one of the main downfalls through our team is that you know, we weren’t keeping that up to date and it was sort of the responsibility of each person who'd asked at the assessment to update that spreadsheet. We we sort of improve things when we actually allocated someone within the team to be in charge. So I think one of the um support work, yeah, sorry the assistants. Physio assistants…and really have assistants, was put in charge of just keeping the spreadsheet up to date, so that just meant that someone was actually chasing the physios to, to input those patients that, that asked, and if they said yes or no, what was the reason?  P18 page 27 line 1080 (physiotherapist) |
| A valuable trial to be involved with | I’m just thrilled to bits I was invited to do it and I just hope that I’ve done everything right you know the more we can do, we can do to help people in the future, it’s amazing. I can’t fault a thing. Everybody's polite kind when they speak to you, explain things not in a patronizing way and those things really matter to me. That nobody sort of looking down their nose or being patronising. It's absolutely lovely, thank you.  P1 page 17 line 536 (participant)  So if you were to do it a similar thing in the future, I'd be happy to do the same.  P12 page 25 line 847 (physiotherapist) |
| The research process works | Wasn't a huge burden. As I said, the amount of it you know additional time on our assessments, there's probably 5-5 minutes max really.  P12 page 24 line 815 (physiotherapist)  Three months to establish a new habit. Potential new habit is about right. I think an hour is really nice because you get a little bit, you get the warm up and the cool down you get a little meditative moment.  P16 page 44 line 1298 (singing leader) |
| Small singing groups | You are thinking why am I the only other person in this singing group? You walk into a big massive room and its just you and the tutor in the room and you spend an hour for what five weeks until one other person comes? It didn’t feel quite good for me.  P23 page 6 line 168 (participant)  I think and if there was a bigger group, it may be better in a lot of instances.  P47 page 51 line 1889 (participant) |
| Home exercises not well adhered to | There was one participant who said I can't. I can't keep up. I can't. I can't fill in all the paperwork. I don't know if I'm doing what I need to do.  P18 page 40 line 1377 (singing leader)  It seems like a lot for jobs for a single lesson…You should understand stepping every day. Yeah, and. You know, if you go, if you go 2 weeks without filling it in, there's nobody to check on you, yeah, and you're basically making it up.  P23 page 18 line 526 (participant) |
| The experience of singing | Pulmonary rehab doesn’t teach you how to breathe. it doesn't teach you actually use your stomach to breathe as the WHAM program. Actually you, you know, you go through, you go through a session, you go through like at at least 10, 10 minute session. And how to do breathing properly, how to breathe directly in your stomach upwards…and so that's that's how the WHAM program is better than the pulmonary program, the pulmonary program never actually teach you how to breathe at all.  P23 page 9 line 259 (participant)  We've just started. Not very long. We are developing a choir…So it's given me, you know, that confidence to actually, you know, because I was told I have a, I have a lovely voice in there. So I've taken that on board.  P31 page 12 line 360 (participant) |
| Social wellbeing an important but missed outcome | The most important benefit was the fact that I met different people, OK, that's I'm, I'm, so there was a social interaction and it's always good for the soul.  P58 page 11 line 448 (participant)  The people in the situation that the people you are you are aiming returning to, are people that have issues with breathing…people that have issue with breathing, issue with going out and issue with self esteem. So basically there's other people are gonna need this type of course. You know, or do the exercise within the course for the rest of your life.  P23 page 5 line 122 (participant) |
| Suggested study improvements | I think the only thing I found a little bit difficult was the case report form. Yeah, you know the case report form it's quite long.  P31 page 10 line 317 (participant)  I think that's unnecessary. And what did you do? And how many times, how did you go? And you know, these are all irrelevant.  P13 page 19 line 639 (participant)  So there probably needs to be a dedicated member of the research team at that site rather than just the clinicians maybe.  P16 page 22 line 643 (singing leader) |

### Table S7: Baseline cohort qualifications

|  | **No. returned** | **Home PR ex** | **SLH + Home PR** | **Difference in proportions** | **P value** | **Confidence Intervals** |
| --- | --- | --- | --- | --- | --- | --- |
| **1 + O level/GCE/GCSE (any grade)** | 15 | 11 | 4 | 0.219 | 0.039 | (.006 - .406) |
| **5 + O level/GCE/GCSE (A-C), School Cerificate** | 18 | 10 | 8 | 0.063 | 0.578 | (-.157 - .275) |
| **1 + AS Level/A Level** | 8 | 5 | 3 | 0.063 | 0.45 | (-.109 - .227) |
| **2 + A Levels/4AS Levels, Higher School Certificate** | 10 | 7 | 3 | 0.125 | 0.168 | (-.061 - .297) |
| **First Degree (BA/BSC)** | 24 | 14 | 10 | 0.125 | 0.302 | (-.112 - .347) |
| **Higher Degree (MA, PhD, PGCE, Post-grad certificates)** | 8 | 4 | 4 | 0 | 1 | (-.168 - .168) |
| **NVQ Level one/Foundation GNVQ** | 1 | 0 | 1 | 0.031 | 0.313 | (-.127 - 0.68) |
| **NVQ Level 2/Intermediate GNVQ** | 1 | 1 | 0 | 0.031 | 0.313 | (-0.68 - .127) |
| **NVQ Level 3/Advanced GNVQ** | 2 | 2 | 0 | 0.063 | 0.151 | (-.052 - .170) |
| **NVQ Level 4-5, HNC, HND** | 0 | 0 | 0 | 0 | NA | (-.080 - 0.80) |
| **Other Quals (i.e City and Guilds,RSA/OCR, BTEC)** | 17 | 8 | 9 | -0.31 | 0.777 | (-.243 - .184) |
| **No Qualifications** | 13 | 4 | 9 | -0.156 | 0.12 | (-.341 - 0.47) |
| **No Professional Qualification** | 19 | 8 | 11 | -0.094 | 0.412 | (-.307 - .130) |
| **Qualified Teacher** | 2 | 1 | 1 | 0 | 1 | (-.112 - .112) |
| **Medical Doctor** | 2 | 1 | 1 | 0 | 1 | (-.112 - .112) |
| **Dentist** | 0 | 0 | 0 | 0 | NA | (-.080 - .080) |
| **Nurse/Midwife/Health Visitor** | 6 | 1 | 5 | -0.125 | 0.086 | (-.268 - .0.33) |
| **Other Professional Qualifications** | 36 | 22 | 14 | 0.25 | 0.044 | (0.006 - .465) |

### Table S8: Baseline Health Economics Data

|  | **No. returned** | **Home PR ex** | **SLH + Home PR** | **Difference in proportions** | **P value** | **Confidence Intervals** |
| --- | --- | --- | --- | --- | --- | --- |
| **Last 3 month hospital attendance for Resp** | 64 | 12 | 4 | 0.25 | 0.021 | (.03 - .44) |
| **Last 3 month hospital attendance for other reasons** | 64 | 16 | 15 | 0.03 | 0.8 | (-.21 - .27) |
|  | **No. returned** | **Home PR Mean (SD)** | **SLH +Home PR Mean (SD)** | **Mean Difference** | **P value** | **Confidence Intervals** |
| **EQ5D5L Descriptive** | 64 | 9.84 (3.41) | 11.44 (3.76) | -1.59 | 0.08 | (-3.39 - .199) |
| **EQ5D5L VAS** | 64 | 68 (18.21) | 65.81 (18.63) | 2.19 | 0.64 | (-7.02 - 11.39) |
| **EQ5D5L Index** | 64 | 0.673 (0.21) | 0.581 (0.26) | 0.09 | 0.11 | (-.24 - .21) |
| **Parking cost (pounds)** | 21 | 0.63 (2.25) | 1.16 (1.77) | -0.54 | 0.63 | (-2.85 - 1.78) |
| **Time out of day for hospital visit (hours)** | 37 | 2.86 (1.52) | 3.33 (2.92) | -0.47 | 0.53 | (-1.96 - 1.02) |
| **Times visited GP at surgery for Resp** | 58 | 0.27 (0.64) | 0.14 (0.45) | 0.12 | 0.4 | (-1.69 - .42) |
| **Times visited GP at surgery for other reasons** | 59 | 0.52 (1.03) | 0.50 (0.79) | 0.02 | 0.947 | (-.47 - .50) |
| **Times with GP at home for Resp** | 58 | 0 | 0 | 0 | 1 | NA |
| **Times with GP at home for other reasons** | 58 | 0 | 0 | 0 | 1 | NA |
| **Times with GP over phone for Resp** | 59 | 0.23(0.82) | 0.24(0.83) | -0.01 | 0.97 | (-.44 - .42) |
| **Times with GP over phone fr other reasons** | 58 | 0.40(1.16) | 0.79(2.06) | -0.39 | 0.38 | (-1.26 - .49) |
| **Times with Nurse at surgery for Resp** | 59 | 0.27(0.45) | 0.07(0.26) | 0.2 | 0.2 | (.01 - .39) |
| **Times with Nurse at surgery for other reasons** | 62 | 0.59(1.72) | 1.23(4.36) | -0.64 | 0.45 | (-2.30 - 1.02) |
| **Times with nurse at home for Resp** | 59 | 0 | 0 | 0 | 1 | NA |
| **Times with nurse at home for other reasons** | 59 | 0.03(0.18) | 0 | 0.03 | 0.33 | (-0.3 - .10) |
| **Times with nurse over phone for Resp** | 60 | 0.17(0.46) | 0.67(0.37) | 0.1 | 0.36 | (-.11 - .31) |
| **Times with nurse over phone for other reasons** | 59 | 0 | 0.14(0.35) | -0.14 | 0.36 | (-.27 - -.01) |
| **Total spent on health product 1 (pounds)** | 36 | 22.95 (47.11) | 17.41(18.01) | 5.54 | 0.65 | (-19.18 - 30.26) |
| **Total spent on health product 2 (pounds)** | 21 | 12.61 (10.21) | 20.67(19.71) | -8.06 | 0.28 | (-23.19 - 7.08) |
| **Days off work or normal duties due to resp** | 55 | 7.12 (14.59) | 5.83 (16.79) | 1.29 | 0.76 | (-7.26 - 9.84) |
| **Days off work or normal duties due to other** | 56 | 2.89 (5.63) | 2.21 (3.76) | 0.68 | 0.6 | (-1.89 - 3.25) |

### Table S9: 12-week follow-up Health Economics data

|  | **No. returned** | **Home PR ex** | **SLH + Home PR** | **Difference in proportions** | **P value** | **Confidence Interval** |
| --- | --- | --- | --- | --- | --- | --- |
| **Last 3 month hospital attendance for Resp** | 59 | 8 | 13 | -0.16 | 0.21 | (-.38 - 0.09) |
| **Last 3 month hospital attendance for other reasons** | 59 | 15 | 16 | -0.02 | 0.8 | (-0.26 - 0.23) |
|  | **No. returned** | **Home PR Mean (SD)** | **SLH +Home PR Mean (SD)** | **Mean Diff** | **P value** | **CI** |
| **EQ5D5L Descriptive** | 59 | 9.31 (3.50) | 11.13(3.55) | -1.82 | 0.052 | (-3.66 - 0.144) |
| **EQ5D5L VAS** | 59 | 64.03 (24.38) | 59.63(23.17) | 4.4 | 0.48 | (-8 - 16.80) |
| **EQ5D5L Index** | 59 | 2.24(8.23) | 3.56 (16.33) | -1.32 | 0.7 | (-8.10 - 5.43) |
| **Parking cost (pounds)** | 31 | 2.07(5.38) | 0.19(0.75) | 1.88 | 0.18 | (-0.90 - 4.65) |
| **Time out of day for hospital visit (hours)** | 39 | 2.96(2.14) | 3.08(2.16) | -0.12 | 0.86 | (-1.53 - 1.28) |
| **Times visited GP at surgery for resp** | 48 | 0.6(1.19) | 1(1.60) | -0.4 | 0.33 | (-1.21 - 0.41) |
| **Times visited GP at surgery for other reasons** | 54 | 0.70(0.91) | 0.44(0.75) | 0.26 | 0.26 | (-0.20 - 0.72) |
| **Times with GP at home for resp** | 59 | 0 | 0.12(0.44) | -0.12 | 0.19 | (-0.3 - 0.62) |
| **Times with GP at home for other reasons** | 48 | 0 | 0 | NA | NA | NA |
| **Times with GP over phone for resp** | 50 | 0.77(0.27) | 0.67(1.31) | -0.59 | 0.029 | (-1.17 - 0.063) |
| **Times with GP over phone for other reasons** | 50 | 0.46(0.93) | 0.46(0.99) | 0 | 0.99 | (-0.55 - 0.54) |
| **Times with Nurse at surgery for resp** | 49 | 0.04(0.20) | 0.29(0.55) | 0.25 | 0.037 | (-.49 - -0.02) |
| **Times with Nurse at surgery for other reasons** | 52 | 0.56(1) | 0.63(0.84) | -0.07 | 0.79 | (-0.58 - 0.44) |
| **Times with nurse at home for resp** | 48 | 0 | 0.13(0.61) | -0.13 | 0.33 | (-0.38 - 0.13) |
| **Times with nurse at home for other reasons** | 48 | 0 | 0 | NA | NA | NA |
| **Times with nurse over phone for resp** | 48 | 0.042 (0.20) | 0.08(0.28) | -0.04 | 0.56 | (-0.19 - 0.10) |
| **Times with nurse over phone for other reasons** | 49 | 0 | 0.04(0.2) | -0.04 | 0.33 | (-.12 - ,04) |
| **Total spent on health product 1 (pounds)** | 41 | 27.41(103.0) | 19.89(37.31) | 7.51 | 0.77 | (-40.83 - 55.86) |
| **Total spent on health product 2 (pounds)** | 25 | 8.03(4.60) | 13.17(14.08) | -5.13 | 0.23 | (-13.65 - 3.39) |
| **Days off work or normal duties due to resp** | 47 | 4.92(9.70) | 17.38(22.57) | -12.46 | 0.014 | (-22.32 - -2.59) |
| **Days off work or normal duties due to other** | 48 | 5.63(9.35) | 6.48(15.87) | 3.67 | 0.82 | (-8.23 - 6.54) |

### Table S10: 6-month Health Economics data

|  | **No. returned** | **Home PR ex** | **SLH + Home PR** | **Difference in proportions** | **P Value** | **Confidence Interval** |
| --- | --- | --- | --- | --- | --- | --- |
| **Last 3 month hospital attendance for resp** | 46 | 6 | 6 | -0.02 | 0.86 | (-0.27 - .23) |
| **Last 3 month hospital attendance for other** | 46 | 13 | 7 | 0.223 | 0.13 | (-.06 - .47) |
|  | **No. returned** | **Home PR Mean (SD)** | **SLH +Home PR Mean (SD)** | **Mean difference** | **P Value** | **Confidence Interval** |
| **EQ5D5L Descriptive** | 44 | 10.68 (4.21) | 11.86(4.12) | -1.18 | 0.352 | (-3.72 - 1.35) |
| **EQ5D5L VAS** | 45 | 64.4 (17.93) | 61.43(22.89 | 3 | 0.625 | (-9.33 - 15.34) |
| **EQ5D5L Index** | 44 | 0.623 (.27) | 0.57(0.24) | 0.05 | 0.5 | (-0.10 - 0.21) |
| **Time out of day for hospital visit (hours)** | 26 | 3.37(1.42) | 3.59(2.27) | -0.22 | 0.759 | (-1.72 - 1.27) |
| **Times visited GP at surgery for resp** | 32 | 0.94(1.71) | 1.13(1.92) | -0.19 | 0.78 | (-1.50 - 1.12) |
| **Times visited GP at surgery for other reasons** | 33 | 0.89(1.05) | 0.86(1.29) | 0.04 | 0.93 | (-0.83 - 0.91) |
| **Times with GP at home for resp** | 29 | 0 | 0 | NA | NA | NA |
| **Times with GP at home for other reasons** | 29 | 0 | 0 | NA | NA | NA |
| **Times with GP over phone for resp** | 32 | 0.38 (0.81) | 1.63(2.90) | -1.25 | 0.11 | (-2.83 - .33) |
| **Times with GP over phone for other reasons** | 30 | 0.38(0.50) | 0.79(1.19) | -0.41 | 0.22 | (-1.08 - 0.31) |
| **Times with Nurse at surgery for resp** | 30 | 0.27(0.46) | 0.40(.63) | -0.13 | 0.51 | (-.55 - 0.28) |
| **Times with Nurse at surgery for other reasons** | 28 | 0.56(1.21) | 0.33(0.49) | 0.23 | 0.54 | (-.53 - 0.99) |
| **Times with nurse at home for resp** | 27 | 0 | 0.08(0.29) | -0.08 | 0.27 | (-0.27 - 0.10) |
| **Times with nurse at home for other reasons** | 27 | 0 | 0 | NA | NA | NA |
| **Times with nurse over phone for resp** | 28 | 0.13(0.5) | 0 | 0.13 | 0.4 | (-0.17 - 0.42) |
| **Times with nurse over phone for other reasons** | 27 | 0.06(0.25) | 0.18(0.40) | -0.12 | 0.35 | (-.41 - .17) |
| **Days off work or normal duties due to resp** | 21 | 7.31(12.89) | 3.13(5.33) | 4.18 | 0.4 | (-5.92 - 14.28) |

### Table S11. Participant demographics for qualitative interviews

| **Participant** | **Sex** | **Age range** | **Respiratory Diagnosis** | **Completion of singing** |
| --- | --- | --- | --- | --- |
| 1 | Female | 65-69 | COPD | Not randomised to singing |
| 4 | Male | 60-64 | COPD | No |
| 7 | Female | 75-79 | COPD | Yes |
| 13 | Male | 80-84 | COPD | Yes |
| 23 | Male | 45-49 | COPD | Yes |
| 28 | Female | 60-64 | Asthma | Yes |
| 31 | Female | 60-64 | Asthma | Yes |
| 36 | Female | 40-44 | ILD | Not randomised to singing |
| 47 | Male | 70-74 | ILD | Yes |
| 58 | Male | 70-74 | COPD | No |

### Table S12: Adverse events

| **Trial ID** | **Total AEs** | **Total SAEs** | **No of non resp AES** | **No of non resp SAES** | **No of resp AES** | **No of resp SAES** |
| --- | --- | --- | --- | --- | --- | --- |
| # | 1 | 1 | 0 | 0 | 1 | 1 |
| # | 1 | 0 | 1 | 0 | 0 | 0 |
| # | 1 | 0 | 1 | 0 | 0 | 0 |
| # | 0 | 1 | 0 | 0 | 0 | 1 |
| # | 2 | 1 | 1 | 1 | 1 | 0 |
| # | 1 | 0 | 0 | 0 | 1 | 0 |
| # | 2 | 0 | 1 | 0 | 1 | 0 |
| # | 1 | 0 | 1 | 0 | 0 | 0 |
| # | 2 | 0 | 1 | 0 | 1 | 0 |
| # | 6 | 0 | 1 | 0 | 5 | 0 |
| # | 3 | 0 | 2 | 0 | 1 | 0 |
| # | 1 | 0 | 0 | 0 | 1 | 0 |
| # | 1 | 0 | 1 | 0 | 0 | 0 |
| # | 1 | 0 | 1 | 0 | 0 | 0 |
| # | 3 | 0 | 2 | 0 | 1 | 0 |
| # | 1 | 2 | 0 | 0 | 1 | 2 |
| # | 0 | 1 | 0 | 1 | 0 | 0 |
| # | 1 | 1 | 0 | 1 | 1 | 0 |
| # | 3 | 0 | 2 | 0 | 1 | 0 |
| # | 3 | 0 | 1 | 0 | 2 | 0 |
| # | 0 | 1 | 0 | 0 | 0 | 1 |
| # | 2 | 0 | 2 | 0 | 0 | 0 |
| # | 4 | 0 | 3 | 0 | 1 | 0 |
| # | 0 | 2 | 0 | 2 | 0 | 0 |
| # | 2 | 2 | 2 | 2 | 0 | 0 |
| # | 2 | 0 | 1 | 0 | 1 | 0 |
| # | 4 | 0 | 1 | 0 | 3 | 0 |
| # | 2 | 0 | 2 | 0 | 0 | 0 |
| # | 1 | 0 | 1 | 0 | 0 | 0 |
| # | 1 | 0 | 1 | 0 | 0 | 0 |
| # | 3 | 0 | 2 | 0 | 1 | 0 |
| # | 1 | 0 | 0 | 0 | 1 | 0 |
| # | 1 | 1 | 0 | 0 | 1 | 1 |
| # | 3 | 0 | 2 | 0 | 1 | 0 |
| # | 2 | 0 | 0 | 0 | 2 | 0 |
| # | 1 | 0 | 1 | 0 | 0 | 0 |
| # | 4 | 0 | 1 | 0 | 3 | 0 |
| # | 0 | 1 | 0 | 1 | 0 | 0 |
| # | 2 | 4 | 2 | 3 | 0 | 1 |
| # | 3 | 0 | 1 | 0 | 2 | 0 |
| # | 2 | 0 | 1 | 0 | 1 | 0 |
| # | 1 | 0 | 0 | 0 | 1 | 0 |
| # | 1 | 0 | 0 | 0 | 1 | 0 |
| # | 2 | 0 | 0 | 0 | 2 | 0 |
| # | 3 | 0 | 2 | 0 | 1 | 0 |
| # | 0 | 4 | 0 | 4 | 0 | 0 |
| # | 3 | 2 | 3 | 2 | 0 | 0 |
| # | 3 | 1 | 3 | 1 | 0 | 0 |
| # | 1 | 0 | 0 | 0 | 1 | 0 |
| # | 2 | 0 | 1 | 0 | 1 | 0 |
| # | 3 | 0 | 2 | 0 | 1 | 0 |
| # | 4 | 0 | 1 | 0 | 3 | 0 |
| # | 2 | 0 | 1 | 0 | 1 | 0 |
| # | 1 | 1 | 0 | 0 | 1 | 1 |
| # | 1 | 0 | 0 | 0 | 1 | 0 |
| # | 1 | 0 | 0 | 0 | 1 | 0 |
| # | 1 | 0 | 1 | 0 | 0 | 0 |
| **Totals** | 103 | 26 | 53 | 18 | 50 | 8 |

### Semi-structured interview guide – Respiratory Participants

Hello. Thank you for offering your time today. I will first summarise the participant information sheet once more and then answer any final questions you may have.

**Summarise PIS, Consent procedure**

How are you today?

Can you give me some of your thoughts on your involvement and experiences on the WHAM trial?

- Were you randomised to singing groups plus PR home exercises or home exercises only?

Was the information you were given about the Trial sufficient in hindsight?

- How could the information be improved?

At what point during your pulmonary rehabilitation did you start to consider participation in the trial?

- What made you want to join the study?

How did you find the singing taster session as part of the PR education programme?

- How would you change it for a future study?
- Where in the education programme would you place the singing taster session if delivered within a cohort programme?
- Did you discuss Singing for Lung Health with clinicians outside of the taster session?

How do you think the process of referral to the research team went?

- In what ways could trial procedures be improved to increase patient recruitment?

Tell me about the assessments performed by the research team.

- Was communication from them acceptable?
- Did the tests and paperwork you were asked to complete make sense?
- How would you change the assessment procedures for a future Trial?

To what extent do the home PR exercise programme and diary/Singing home exercise diary need to be refined or adapted to make it more relevant to you?

- To what extent do you think the amount of combined singing and PR home exercises is acceptable?
- How can we improve completion rates of these diaries?

What was your experience like participating in Singing for Lung Health groups/ PR home exercises during the Trial?

- Covid-19 considerations
- Liaison with research staff (did you feel adequately supported?)
- Liaison with singing leader
- Is the dosing (frequency, length of sessions etc) acceptable?

How could the delivery of the singing groups or home exercises be refined or adapted to make it acceptable, relevant or useful for you?

How do you think Singing for Lung Health maintains benefits gained from PR (If randomised to singing groups)?

What options were given to you at the end of the trial about joining singing for lung health groups?

Did your involvement in the Trial have any unanticipated negative impacts?

How would you change how the study has been designed?

Any other comments?

Thank you for your time again. We will be in contact regarding the results of the study
